# Metabolic profiling and longitudinal changes in patients with severe aortic stenosis undergoing valve replacement

**DOI:** 10.1101/2025.08.01.25332646

**Authors:** Christian Nitsche, George Thornton, Jonathan Bennett, Francisco Gama, Suchi Chadalavadha, Nish Chaturvedi, Therese Tillin, Charlotte Manisty, Arantxa González, Eylem Levelt, James C Moon, Alun D Hughes, Anish N Bhuva, Thomas A Treibel

## Abstract

**Aims:** In patients with aortic stenosis (AS), the relation of cardiac energetic pathways with cardiac structure and function, their changes, and their prognostic significance are not well understood. We aimed to characterise metabolic profiles in patients with severe AS before and after aortic valve replacement (AVR) and their association with functional status, structural remodelling and mortality.

**Methods and results:** Patients with symptomatic, severe AS before (n=143) and 1-year after (n=113) AVR underwent cardiac magnetic resonance (CMR), serum cardiac biomarkers, and 6-minute walk test. Resting non-fasting plasma samples underwent targeted nuclear magnetic resonance for free fatty acids (FFAs), branched chain amino acids (BCAAs), glycolysis-related metabolites, and ketones.

Lower FFAs and BCAA concentrations, but not glycolysis metabolites or ketones, correlated with greater myocardial mass and focal fibrosis, NT-proBNP, TnT and 6-minute walk distance. After 10.5 years of follow-up (66/143 deaths), lower FFAs and BCAAs, but not ketones were independently associated with higher mortality risk (*P*<0.05). At 1-year after AVR, FFAs had decreased compared to baseline.

**Conclusion:** Reduced serum FFAs and BCAA concentrations are cardiac, maladaptive metabolic changes to AS, and are not reversible after AVR. Whether these markers may be used to guide the timing of AVR or may be targeted by metabolic interventions remains to be evaluated by future research.

**Condensed Abstract:** In patients with aortic stenosis (AS) systemic metabolomics and their association with myocardial remodeling and outcome after aortic valve replacement (AVR) are largely unknown. We show that in severe AS low levels of unsaturated fatty acids and branch chain amino acids correlate with worse disease markers by cardiac magnetic resonance, biomarkers, and functional incapacity, do not increase after AVR and correlate with higher mortality. Targeted manipulation of metabolite perturbations might ameliorate myocardial energetics before and after AVR.

**Graphical abstract:** 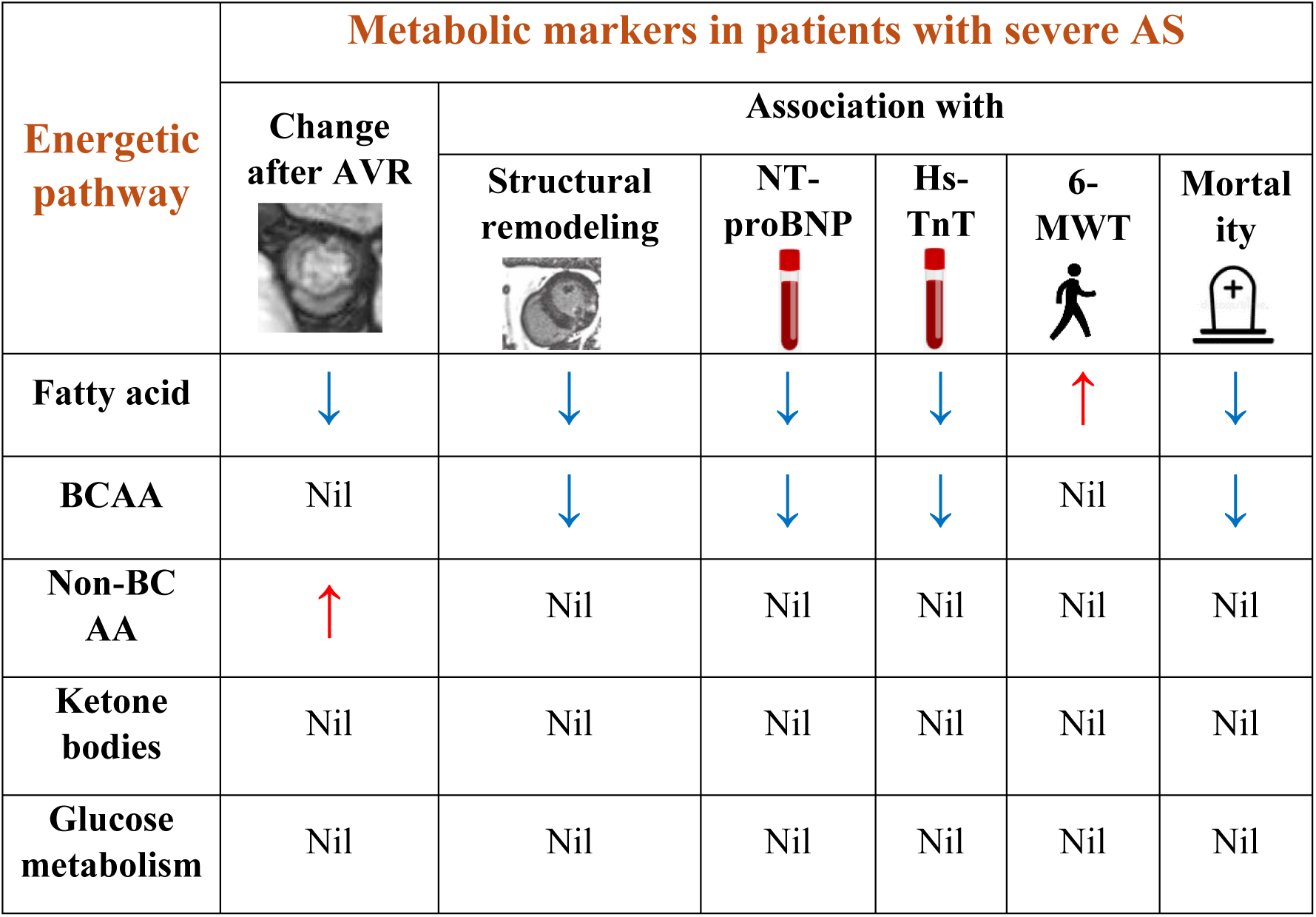

Metabolites in severe aortic stenosis and their associations with cardiac remodeling/damage and mortality after aortic valve replacement (AVR). FFAs indicates free fatty acids; BCAA, branched chained amino acids;

## Introduction

In aortic stenosis (AS) progressive narrowing of the aortic valve causes pressure overload of the left ventricle, myocardial remodeling and heart failure. Structural alterations of the myocardium are accompanied by changes in myocardial energy metabolism. Whereas the normal heart predominantly uses fatty acids (FA) as energy source (60 to 90%), ^1, 2^ substrate utilisation is altered in AS with less FA oxidation and increased glucose use. ^3, 4^ It is unclear whether changes in myocardial metabolism are adaptive or maladaptive, and whether they are reversible. High throughput metabolomic profiling permits systematic assessment of metabolomic pathways related to the heart. ^5, 6^ Serum changes may reflect systemic adaptation to the haemodynamic consequences of AS, or intrinsic myocardial metabolic remodeling. Linkage of metabolic alterations to functional, structural, and biochemical markers of myocardial injury and their trajectories after intervention may help to untangle this relationship.

A more nuanced understanding of metabolic derangement in AS may be of substantial clinical interest as therapeutic strategies aimed at manipulation of cardiac substrate utilisation have not proven to be of substantial benefit, and untangling the interplay between cardiac structural changes and metabolism may yield pathophysiological insights. ^2^

We hypothesised that plasma metabolic markers in AS would: 1) associate with imaging, biochemical and functional cardiovascular disease markers, 2) normalise after valve replacement and 3) portend prognostic value.

## Methods

### Study population

In a prospective observational cohort study, patients with severe symptomatic AS who underwent AVR between January 2012 and January 2015 were included at a single tertiary referral cardiac center, University College London Hospital NHS Trust, London, United Kingdom. The study was approved by the ethical committee of the U.K. National Research Ethics Service (07/H0715/101) and this work is a substudy of the previously published RELIEF-AS study (Regression of Myocardial Fibrosis After Aortic Valve Replacement; NCT02174471). ^7^ The study conformed to the principles of the Helsinki Declaration, and all subjects gave written informed consent. Patients or the public were not involved in the design, or conduct, or reporting, or dissemination plans of our research. Patients were recruited before pre-operative evaluation. Pre-AVR and post-AVR, a comprehensive assessment included clinical history, blood pressure, 6-min walk test (6MWT), blood sampling (for high-sensitivity troponin T [hsTnT] and N-terminal pro–B-type natriuretic peptide [NT-proBNP]), electrocardiography, trans-thoracic echocardiography, and cardiac magnetic resonance (CMR) using the same equipment. Non fasting blood was collected, and plasma was separated by centrifugation and stored at-80 degrees Celsius. Inclusion and exclusion criteria for the study have previously been described. ^7^ In short, only patients with severe AS and clear indication for AVR, no greater than moderate valve disease other than AS, and ability to undergo CMR were included.

### Study procedures

#### Metabolic marker profiling

A targeted nuclear magnetic resonance (NMR) metabolomics platform was used for the quantification of metabolites in serum samples. ^6, 8–10^ Metabolite assessment was performed at baseline (pre-AVR) and follow-up (at 1-year after AVR). Overall, out of >150 metabolites measured, 21 metabolites were selected as reflecting the main energetic pathways related to the heart: FAs, amino acids, glucose, lactate, and ketone bodies. ^11^ FA composition included saturated, monounsaturated (MUFA) and polyunsaturated (PUFA) FAs. Glycolytic metabolites (glucose, citrate) were measured in relation to glucose metabolisms. Amino acids (AAs) were categorized into branched-chained AAs, levels of which have been shown to undergo changes in pressure-overloaded mouse models, ^12^ and non-branched-chained AAs. Acetate and 3-hydroxybutyrate were measured as ketone bodies. The reliability of assays was assessed in a sample of 12 patients who underwent repeat analysis on a second serum sample. This showed excellent concordance (intra class correlation coefficient 0.92-0.99 for all metabolites analysed).

#### Multimodality cardiac imaging

Echocardiography was primarily used to assess ventricular function, diastolic parameters and valve area or velocities. Contrast-enhanced CMR was performed for deep structural and functional phenotyping as previously reported, ^7^ and included cine imaging, quantitative late gadolinium enhancement (LGE), T1 mapping and extracellular volume fraction. LGE was quantified with the 3-standard deviation above normal myocardium method and calculated as percentage of the LV. All analysis were performed by operators blinded to clinical parameters. CMR image analysis was done using CVI42 software (version 5.1.2[303], Circle Cardiovascular Imaging, Calgary, Alberta, Canada).

#### Outcome analysis

All-cause mortality was captured from the National Health Service Spine database and was 100% complete.

## Statistical analysis

All continuous variables are expressed as mean±standard deviation (SD) or median (interquartile range [IQR]) for skewed data. Normality was checked using the Shapiro-Wilk test. Categorical variables are expressed as percentages. Groups were compared using the Mann-Whitney U test or test chi-square test, as appropriate. Changes between pre-AVR and post-AVR visits were compared using the paired t-test and McNemar’s test, as appropriate.

To identify associations with quantitative markers of cardiac structure (LVMi), cardiac damage (TnT, NT-proBNP, extent of focal scar on CMR), and functional capacity (6-MWT) and their longitudinal changes, multiple linear regression analysis was performed for all metabolomic markers. Adjustment of baseline models was performed for potential confounders (age, sex, BMI, arterial hypertension, diabetes), which were chosen based on prior knowledge. A false discovery rate of 0.1 was used to determine significant associations. To test whether baseline serum metabolites explain cardiac reverse remodeling and functional recovery (i.e., the change in TnT, NT-proBNP, 6-MWT), regression models were adjusted for age, sex, BMI, arterial hypertension, diabetes, delta mean transvalvular gradient, and the baseline value of the respective marker. The association of metabolite components with all-cause mortality was assessed using multivariate Cox proportional hazard regression models with adjustment performed for potential confounders (age, sex, hypertension, diabetes). Univariate models are also presented. A two-sided p-value of <0.05 was considered significant. Statistical analyses were carried out using SPSS software version 28 (IBM, Armonk, New York).

## Results

### Patient characteristics

In total, 181 people with severe AS were screened and 38 were excluded due to various reasons (**Figure 1**). The remaining 143 patients with both metabolomic and multimodality imaging data formed the baseline AS cohort. Of these 143 AS patients, 113 returned for 1-year follow-up with repeat metabolomic and imaging (n=106, post-AVR pacemaker in 7 patients with only metabolomic evaluation at follow-up) assessment. Detailed patient characteristics are displayed in **Tables 1 and 2**.

**Figure 1.**
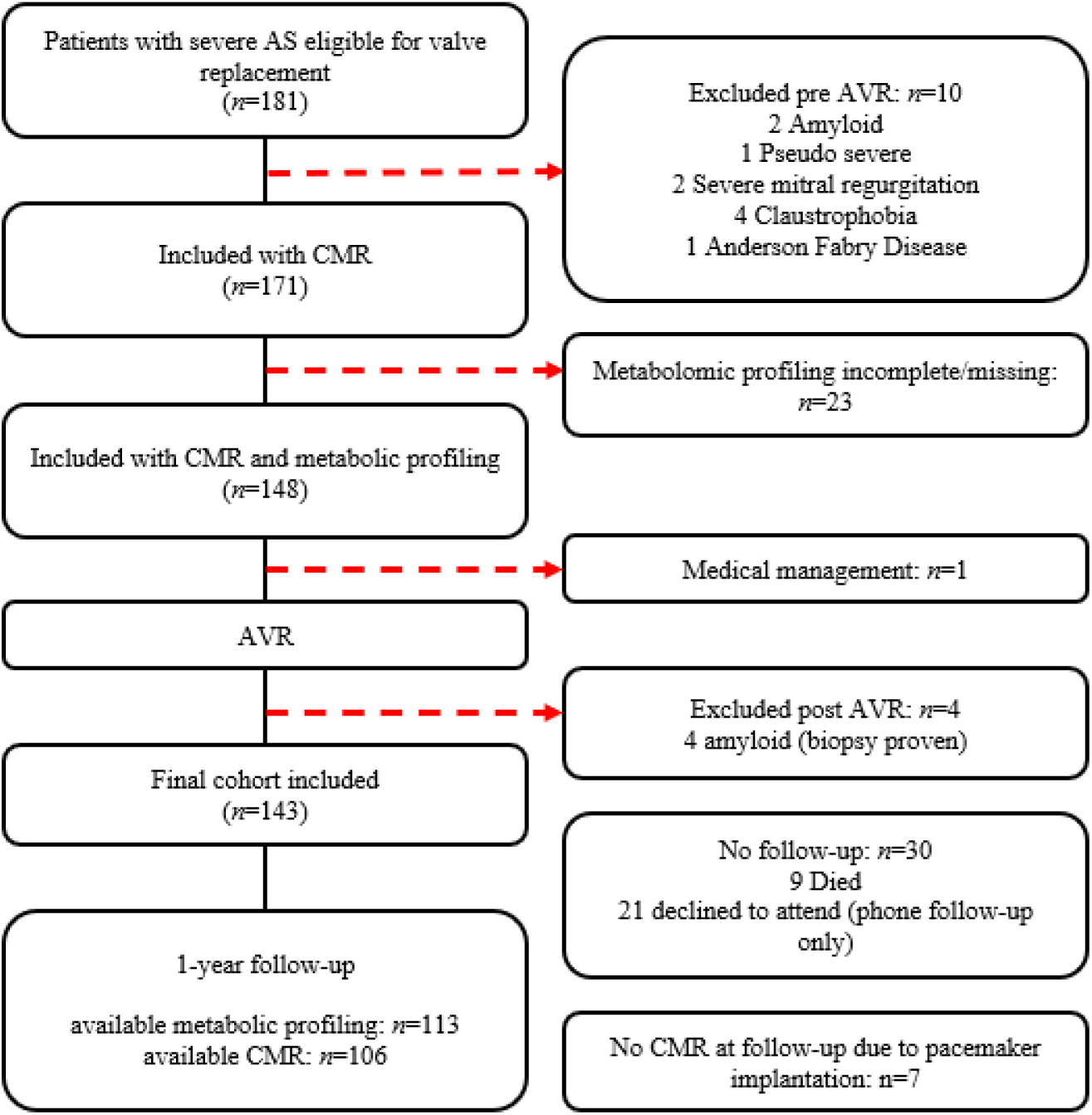
Patient flow chart. AS indicates aortic stenosis; CMR, cardiac magnetic resonance imaging; AVR, aortic valve replacement;

**Table 1.** Patient baseline characteristics.

|  | Severe Aortic Stenosis<br>*n*=143 |
| --- | --- |
| *Demographics* |  |
| Age, years | 72 (65-77) |
| Male sex, % | 55.2 |
| BMI, kg/m<sup>2</sup> | 28.0 (24.9-31.5) |
| Ethnicity, % |  |
| European | 93.8 |
| South Asian | 4.7 |
| African Caribbean | 1.6 |
| EuroSCORE-II, % | 1.5 (1.0-2.5) |
| *Clinical parameters* |  |
| Angina, % | 36.6 |
| Syncope, % | 9.2 |
| NYHA class III+, % | 37.5 |
| 6-minute walk distance, m | 480 (333-584) |
| Diabetes, % | 19.7 |
| Hypertension, % | 80.3 |
| Atrial fibrillation, % | 16.1 |
| Coronary artery disease, % | 31.0 |
| *Laboratory results* |  |
| NT-proBNP, pg/ml | 76 (30-251) |
| Troponin, pg/ml | 14 (10-20) |
| eGFR, ml/min | 73 (60-92) |
| *Imaging parameters* |  |
| *Echocardiography* |  |
| Mean gradient, mmHg | 46 (38-55) |
| *CMR* |  |
| Vmax, m/s | 3.8 (3.3-4.2) |
| LVEF, % | 73 (64-80) |
| Stroke volume, ml | 82 (69-95) |
| LV mass index, g/m<sup>2</sup> | 83 (70-106) |
| LGE mass*, % | 9.2 (5.0-17.5) |

Continuous parameters are shown as median and interquartile ranges and categorical as percentages.
\*) LGE mass (percent) as the median of all patients, including those without LGE.
BMI indicates body mass index; NYHA, New York Heart Association functional class; NT-proBNP, N-terminal pro-brain natriuretic peptide; eGFR, estimated glomerular filtration rate; Vmax, maximum transaortic velocity; LVEF, left ventricular ejection fraction; LGE, late gadolinium enhancement.

**Table 2.** Baseline metabolomics.

|  | Severe Aortic Stenosis<br>*n*=143 |
| --- | --- |
| <u>Glucose metabolism</u> |  |
| Glucose, mmol/l | 4.9 (4.1-6.1) |
| Lactate, mmol/l | 1.2 (1.0-1.6) |
| Citrate, mmol/l | 0.19 (0.17-0.21) |
| <u>Fatty acids</u> |  |
| Total fatty acids, mmol/l | 10.8 (8.8-12.6) |
| Estimated degree of unsaturation | 1.24 (1.19-1.28) |
| Omega-3 fatty acids, mmol/l | 4.1 (3.1-5.1) |
| 22:6, docosahexaenoic acid, mmol/l | 0.13 (0.10-0.17) |
| Omega-6 fatty acids, mmol/l | 3.4 (2.8-4.0) |
| 18:2, linoleic acid, mmol/l | 2.8 (2.2-3.3) |
| Polyunsaturated fatty acids, mmol/l | 3.9 (3.1-4.5) |
| Monounsaturated fatty acids; 16:1, 18:1,<br>mmol/l | 2.9 (2.2-3.4) |
| Saturated fatty acids, mmol/l | 4.2 (3.5-4.7) |
| <u>Amino acids (AA)</u> |  |
| *Non-branched chained AA* |  |
| Alanine, mmol/l | 0.40 (0.36-0.45) |
| Glutamine, mmol/l | 0.46 (0.40-0.53) |
| Histidine, mmol/l | 0.05 (0.05-0.06) |
| *Branched chained AA* |  |
| Isoleucine, mmol/l | 0.06 (0.05-0.08) |
| Leucine, mmol/l | 0.08 (0.07-0.10) |
| Valine, mmol/l | 0.16 (0.13-0.18) |
| *Aromatic AA* |  |
| Phenylalanine, mmol/l | 0.06 (0.06-0.07) |
| Tyrosine, mmol/l | 0.06 (0.05-0.07) |
| <u>Ketone bodies</u> |  |
| Acetoacetate, mmol/l | 0.05 (0.05-0.06) |
| 3-hydroxybutyrate, mmol/l | 0.11 (0.10-0.14) |

### Association of metabolic markers and cardiac structure, functional capacity, troponin-T and NT-proBNP

Full univariate and multivariate models are presented in the supplement (**Table S1**). Lower FA levels were independently associated with more LV remodeling, scar, and damage, and worse functional capacity as shown by higher LVMi, higher amount of LGE, higher NT-proBNP and hs-TnT, and lower 6-minute walk distance. Lower levels of branched-chained AAs were linked to higher cardiac damage, as indicated by significant associations of Isoleucin and Leucin with LVMi (P=0.001 and 0.002), NT-proBNP (both P<0.001) and hs-TnT (P=0.024 and 0.013). Also, valine was significantly associated with NT-proBNP (P=0.002) and hs-TnT (P=0.016).

### Changes in metabolic markers after AVR

At 1-year after AVR, symptoms, valve gradients, structural LV remodeling, and markers of cardiac injury and decompensation improved (**Table 3**). Levels of FA in AS had decreased at 1-year post-AVR (e.g., Omega-6 FA: 3.5 to 3.2 mmol/l; Omega-3 FA: 4.1 to 3.9 mmol/l, both *P*<0.05). AA and ketone body concentrations had not changed at follow-up (all *P*>0.05).

**Table 3.**
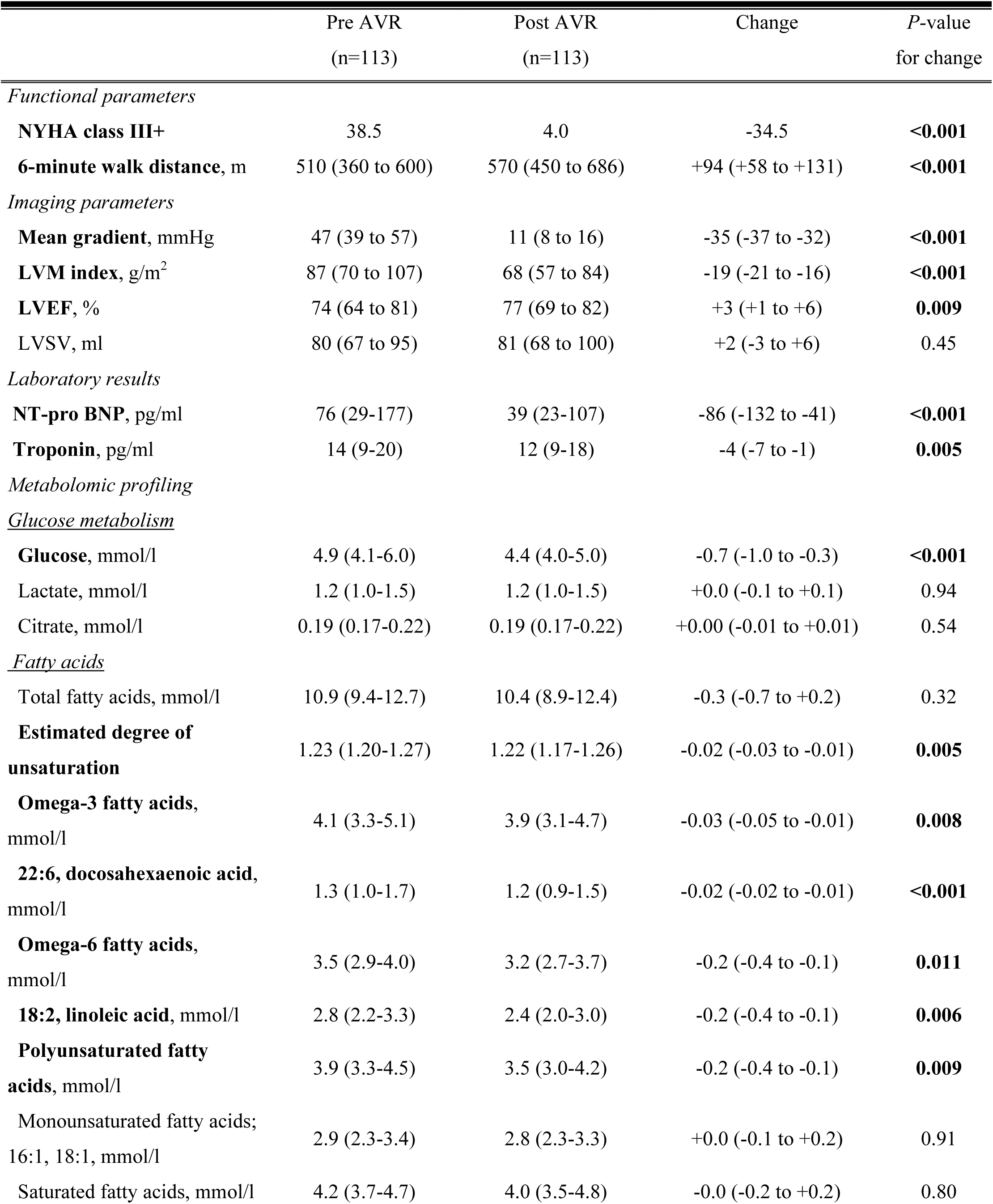
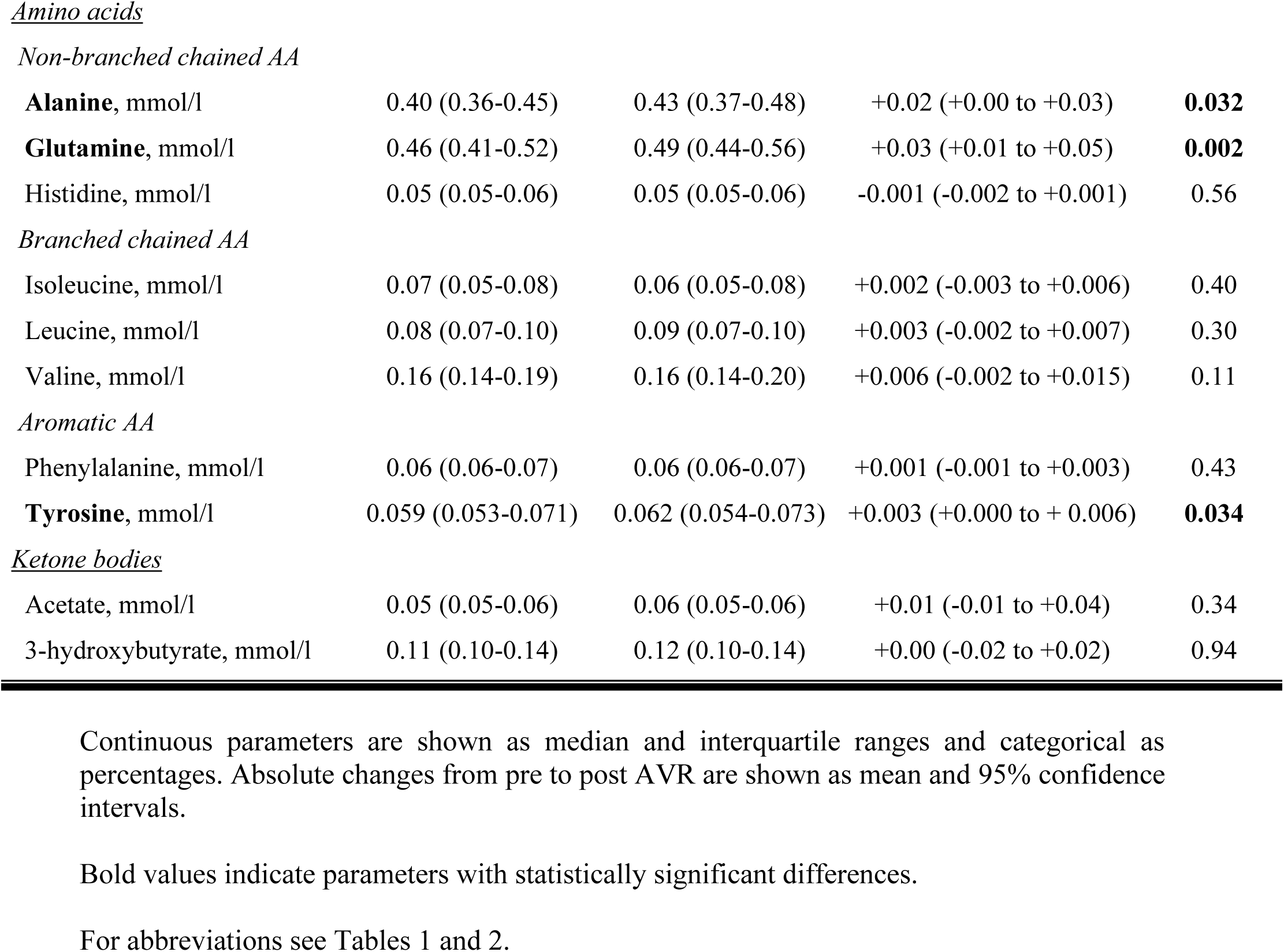
Changes in endpoints before and after AVR.

|  | Pre AVR<br>(n=113) | Post AVR<br>(n=113) | Change | P-value<br>for change |
| --- | --- | --- | --- | --- |
| *Functional parameters* |  |  |  |  |
| **NYHA class III+** | 38.5 | 4.0 | -34.5 | **<0.001** |
| **6-minute walk distance, m** | 510 (360 to 600) | 570 (450 to 686) | +94 (+58 to +131) | **<0.001** |
| *Imaging parameters* |  |  |  |  |
| **Mean gradient, mmHg** | 47 (39 to 57) | 11 (8 to 16) | -35 (-37 to -32) | **<0.001** |
| **LVM index, g/m<sup>2</sup>** | 87 (70 to 107) | 68 (57 to 84) | -19 (-21 to -16) | **<0.001** |
| **LVEF, %** | 74 (64 to 81) | 77 (69 to 82) | +3 (+1 to +6) | **0.009** |
| **LVSv, ml** | 80 (67 to 95) | 81 (68 to 100) | +2 (-3 to +6) | 0.45 |
| *Laboratory results* |  |  |  |  |
| **NT-pro BNP, pg/ml** | 76 (29-177) | 39 (23-107) | -86 (-132 to -41) | **<0.001** |
| **Troponin, pg/ml** | 14 (9-20) | 12 (9-18) | -4 (-7 to -1) | **0.005** |
| *Metabolomic profiling* |  |  |  |  |
| <u>*Glucose metabolism*</u> |  |  |  |  |
| **Glucose, mmol/l** | 4.9 (4.1-6.0) | 4.4 (4.0-5.0) | -0.7 (-1.0 to -0.3) | **<0.001** |
| **Lactate, mmol/l** | 1.2 (1.0-1.5) | 1.2 (1.0-1.5) | +0.0 (-0.1 to +0.1) | 0.94 |
| **Citrate, mmol/l** | 0.19 (0.17-0.22) | 0.19 (0.17-0.22) | +0.00 (-0.01 to +0.01) | 0.54 |
| <u>*Fatty acids*</u> |  |  |  |  |
| **Total fatty acids, mmol/l** | 10.9 (9.4-12.7) | 10.4 (8.9-12.4) | -0.3 (-0.7 to +0.2) | 0.32 |
| **Estimated degree of unsaturation** | 1.23 (1.20-1.27) | 1.22 (1.17-1.26) | -0.02 (-0.03 to -0.01) | **0.005** |
| **Omega-3 fatty acids, mmol/l** | 4.1 (3.3-5.1) | 3.9 (3.1-4.7) | -0.03 (-0.05 to -0.01) | **0.008** |
| **22:6, docosahexaenoic acid, mmol/l** | 1.3 (1.0-1.7) | 1.2 (0.9-1.5) | -0.02 (-0.02 to -0.01) | **<0.001** |
| **Omega-6 fatty acids, mmol/l** | 3.5 (2.9-4.0) | 3.2 (2.7-3.7) | -0.2 (-0.4 to -0.1) | **0.011** |
| **18:2, linoleic acid, mmol/l** | 2.8 (2.2-3.3) | 2.4 (2.0-3.0) | -0.2 (-0.4 to -0.1) | **0.006** |
| **Polyunsaturated fatty acids, mmol/l** | 3.9 (3.3-4.5) | 3.5 (3.0-4.2) | -0.2 (-0.4 to -0.1) | **0.009** |
| **Monounsaturated fatty acids; 16:1, 18:1, mmol/l** | 2.9 (2.3-3.4) | 2.8 (2.3-3.3) | +0.0 (-0.1 to +0.2) | 0.91 |
| **Saturated fatty acids, mmol/l** | 4.2 (3.7-4.7) | 4.0 (3.5-4.8) | -0.0 (-0.2 to +0.2) | 0.80 |
| *Non-branched chained AA* |  |  |  |  |
| **Alanine**, mmol/l | 0.40 (0.36-0.45) | 0.43 (0.37-0.48) | +0.02 (+0.00 to +0.03) | **0.032** |
| **Glutamine**, mmol/l | 0.46 (0.41-0.52) | 0.49 (0.44-0.56) | +0.03 (+0.01 to +0.05) | **0.002** |
| Histidine, mmol/l | 0.05 (0.05-0.06) | 0.05 (0.05-0.06) | -0.001 (-0.002 to +0.001) | 0.56 |
| *Branched chained AA* |  |  |  |  |
| Isoleucine, mmol/l | 0.07 (0.05-0.08) | 0.06 (0.05-0.08) | +0.002 (-0.003 to +0.006) | 0.40 |
| Leucine, mmol/l | 0.08 (0.07-0.10) | 0.09 (0.07-0.10) | +0.003 (-0.002 to +0.007) | 0.30 |
| Valine, mmol/l | 0.16 (0.14-0.19) | 0.16 (0.14-0.20) | +0.006 (-0.002 to +0.015) | 0.11 |
| *Aromatic AA* |  |  |  |  |
| Phenylalanine, mmol/l | 0.06 (0.06-0.07) | 0.06 (0.06-0.07) | +0.001 (-0.001 to +0.003) | 0.43 |
| **Tyrosine**, mmol/l | 0.059 (0.053-0.071) | 0.062 (0.054-0.073) | +0.003 (+0.000 to +0.006) | **0.034** |
| Ketone bodies |  |  |  |  |
| Acetate, mmol/l | 0.05 (0.05-0.06) | 0.06 (0.05-0.06) | +0.01 (-0.01 to +0.04) | 0.34 |
| 3-hydroxybutyrate, mmol/l | 0.11 (0.10-0.14) | 0.12 (0.10-0.14) | +0.00 (-0.02 to +0.02) | 0.94 |

- - 1 - 2 Continuous parameters are shown as median and interquartile ranges and categorical as - 3 percentages. Absolute changes from pre to post AVR are shown as mean and 95% confidence - 4 intervals. - 5 - 6 Bold values indicate parameters with statistically significant differences. - 7 - 8 For abbreviations see Tables 1 and 2.

### Association of metabolic markers and changes in cardiac structure, functional capacity, Troponin-T and NT-proBNP after AVR

Detailed analysis of associations of metabolomics with changes in cardiac damage markers after AVR are displayed in **Tables S2 and S3**. Among baseline metabolic markers independently associated with the longitudinal regression of LVMi were lower levels of PUFAs and ketone bodies and higher levels of non-branched-chained AA. Interestingly, even though branched-chained AA did not undergo significant changes from pre-to post-AVR in the overall cohort, their longitudinal increase was associated with a higher decline in hs-TnT [ΔIsoleucine: ß=-0.11 (-0.20 to-0.02); ΔLeucine: ß=-0.11 (-0.19 to-0.02); ΔValine: ß= - 0.11 (-0.20 to-0.03); P for all<0.05].

### Prognostic value of metabolic markers

After 10.5 (IQR 9.9-10.9) years of follow-up, 46% (n=66/143) of patients with severe AS and successful AVR had died. Details of associations of metabolic markers with outcome on uni-and multivariate analysis are displayed in **Table 4**. On univariate analysis, metabolites associated with improved survival post-AVR were higher omega-3 FA and higher branched-chained AA levels. After adjustment for age and sex, diabetes and hypertension, significant associations with lower mortality persisted for higher omega-3 FA levels (adjHR 0.71, 95% CI 0.53-0.95, P=0.02), and higher levels of branched-chained AA (Isoleucin: adjHR 0.72, 95% CI 0.54-0.96; Leucin: adjHR 0.67, 95% CI 0.50-0.90; Valine: adjHR 0.63, 95% CI 0.47-0.84; P<0.05 for all; **Figure 2**). Ketone bodies were not associated with survival.

**Figure 2.**
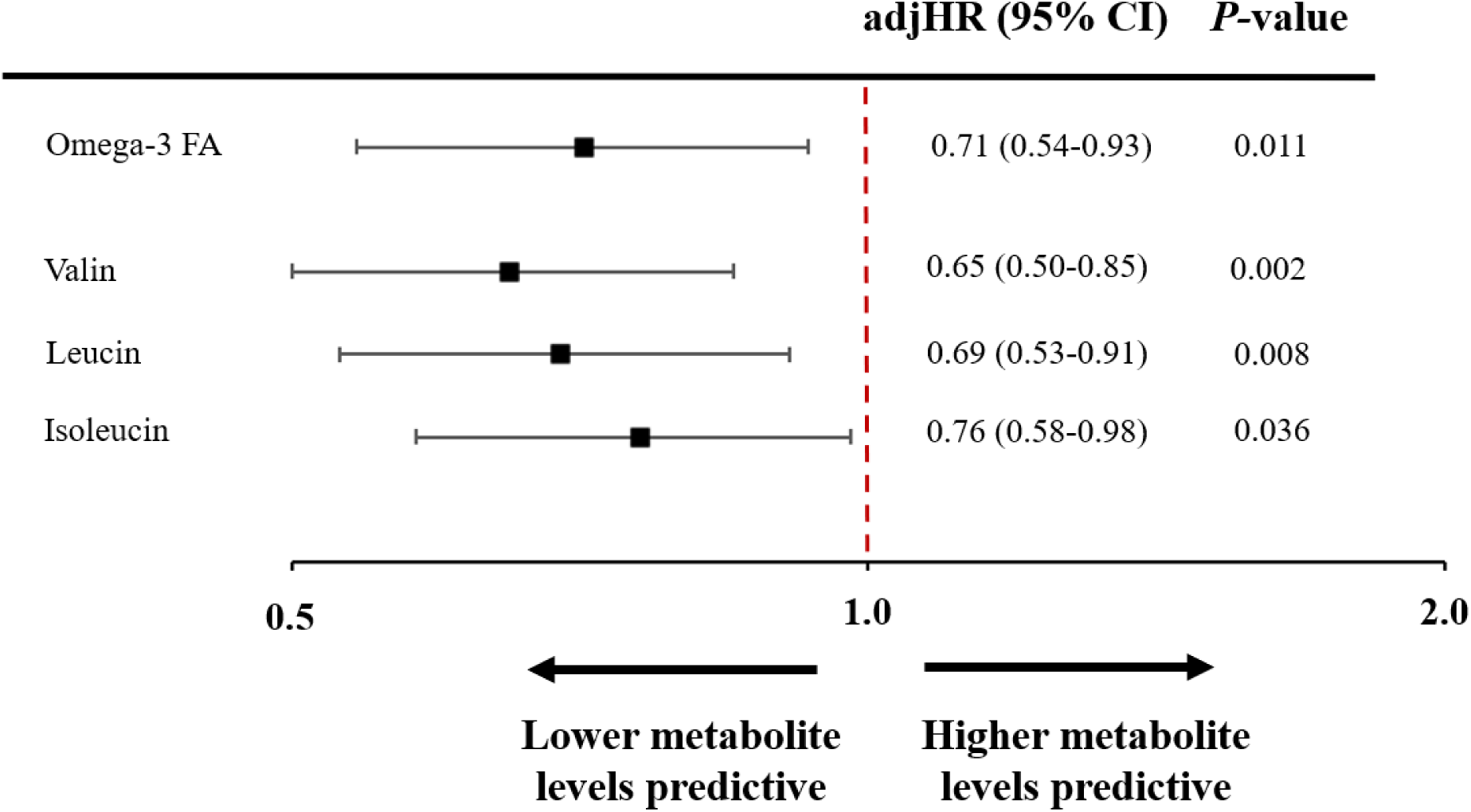
Survival impact of metabolic markers. Higher levels of omega-3 fatty acids, and branched chained amino acids (Valin, Leucin, Isoleucin) were associated with reduced adjusted risk of mortality. Other fatty and amino acids, and ketone bodies showed no association with mortality. AdjHR indicates adjusted hazard ratio; CI, confidence interval;

**Table 4.**
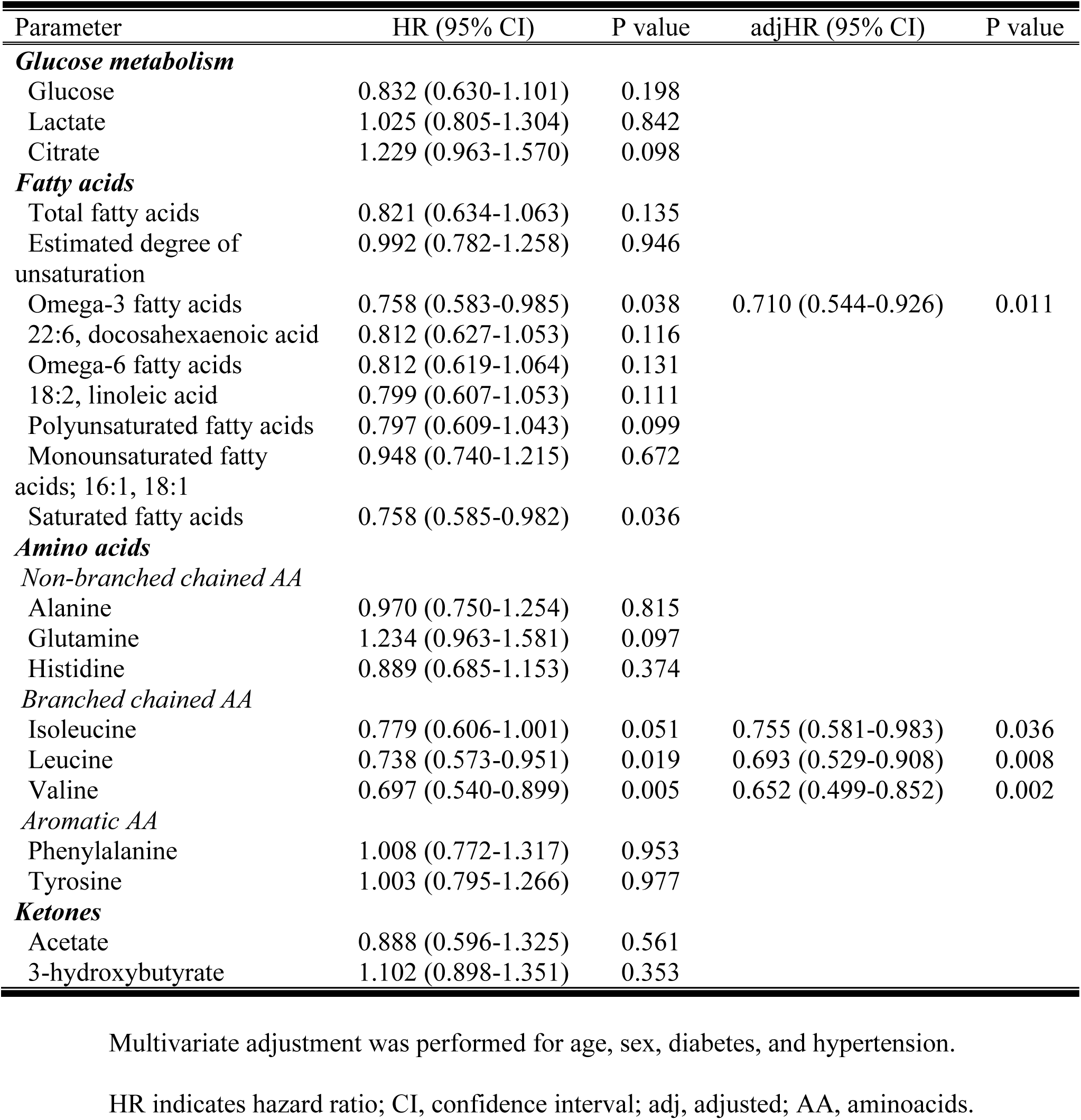
Outcome analysis assessing the association of metabolite components with the risk of mortality.

## Discussion

In severe AS, the metabolic milieu undergoes substantial changes that can be captured with systemic metabolic profiling. The present study assessed plasma metabolic markers coupled with deep structural phenotyping by CMR in severe AS before and after valve replacement and linked these markers with long-term clinical outcome. Lower fatty acid and branched-chained amino acid levels were related to adverse LV remodeling, scar, strain, and poor prognosis and therefore appear to be maladaptive in AS (**Graphical abstract**). Valve replacement did not result in an increase of fatty acid and amino acid concentrations at one year follow-up.

This study is the first in severe AS to associate metabolic signatures with deep structural phenotyping by CMR, which is considered the gold standard for the assessment of cardiac structure, function, and tissue characterisation. Blood metabolic signatures in AS have previously been linked to maladaptive LV remodeling and strain by echocardiography. ^13, 14^ These associations may represent myocardial metabolic alterations or direct/indirect hemodynamic sequalae of AS. In the present study, lower FA levels correlated with features of adverse LV remodeling and elevated cardiac biomarkers. The cascade of cardiac damage in AS may entail fluid retention and intestinal congestion. ^15–17^ Polyunsaturated fatty acids (PUFAs) reflect dietary biomarkers of fatty acid intake and intestinal absorption which may be significantly impeded in the setting of systemic congestion. Decreased systemic perfusion as a result from forward failure in the setting of severe AS may aggravate this condition. Fatty acid levels did not increase at one year post AVR. These observations coupled with adverse prognostic implications post-AVR indicate that a decrease in FA levels is maladaptive and irreversible in severe symptomatic AS.

Similarly, we describe for the first time that lower levels of branched-chained AAs (BCAA) were linked to cardiac biomarker elevation and more pronounced LV remodeling.

Decreased levels of BCAAs have previously been reported in AS compared to controls. ^14^ This finding may be due to the shift of cardiac substrate utilization with increased BCAA consumption in the setting of AS and/or due to kidney dysfunction, which is prevalent in AS with associated heart failure. Patients with a longitudinal increase in BCAA levels displayed a higher decline in NT-proBNP and hs-TnT, indicating that BCAA metabolism may be plastic in a subset of patients and linked to cardiac strain recovery after valve replacement. Importantly, low pre-interventional BCAA levels were independently associated with unfavorable survival post-AVR. These findings suggest that low BCAA levels represent a high-risk marker in severe AS.

### Informing potential future therapeutic interventions

Results from the present study may support the use of therapeutic interventions aimed at manipulating the metabolic profiles in AS to a) maintain a beneficial myocardial metabolic state, and to b) enable/accelerate normalisation of metabolic perturbations following valve replacement. SGLT-2 inhibitors are believed to improve cardiovascular outcomes through pleiotropic effects including a shift in myocardial fuel utilization patterns (increased ketone and FA consumption). ^18^ Other agents, like GLP-1 receptor agonists, likely have no effect on cardiac substrate use, but improve cardiac function through systemic as well as other myocyte effects. ^19^ The complex interplay between substrate availability/utilization, loading conditions and LV remodeling/function highlights the unmet need to assess the efficacy of pharmaceutical interventions with metabolic effects in AS.

### Study limitations

Several limitations of the present study merit comment. Presented data were collected in a single center setting. Therefore, a center-specific bias cannot be excluded. However, the major advantages of limiting data collection to a single center are (i) inclusion of a homogenous patient population, (ii) adherence to a constant clinical routine, and (iii) constant quality of CMR work-up. Also, cardiac metabolite concentrations were not measured directly within myocardial tissue. A higher serum concentration is an indirect evaluation of cardiac metabolism but the rate of FA, ketone body and amino acid uptake into the myocardium is linearly associated with the plasma concentration and changes correlated with other detailed cardiac phenotypes. ^20^

## Conclusions

Linking metabolites with cardiac biomarkers and deep structural phenotyping by CMR revealed significant associations of low FA and BCAA levels with markers of more pronounced LV remodeling and cardiac strain/damage. Low FA and BCAA levels were linked to poor outcome post-AVR rendering them high-risk markers in severe AS. In summary, our data shows that blood metabolite profiles in severe AS go beyond linkage to myocardial structure but portend prognostic value. Whether these markers may be used to guide the timing of AVR or may be targeted by metabolic interventions remains to be evaluated by future research.

## Abbreviations

AA: Amino acids
AS: Aortic stenosis
AVR: Aortic valve replacement
BCAA: Branched-chained amino acids
CMR: Cardiac magnetic resonance
FA: Fatty acids
LGE: Late gadolinium enhancement
MUFA: Monounsaturated fatty acids
PUFA: Polyunsaturated fatty acids

## Data Availability

Data will be available upon reasonable request to the corresponding author

